# Prostate-Specific Membrane Antigen PET/CT Complements MRI for Focal Therapy Selection in Prostate Cancer: A Whole-Mount Histopathologic Study

**DOI:** 10.64898/2026.09.09.26362553

**Authors:** Ida Sonni, Zahra Mohammadigoldar, George Furey, Koichiro Kimura, Pranav C. Movva, Adrien Holzgreve, Shadfar Bahri, Tyler Sevco, Alex Chung, Anthony Sisk, Tristan Grogan, Johannes Czernin, Kyunghyun Sung, Jeremie Calais, Scott E. Eggener, Wayne Brisbane, Robert E. Reiter, Steven Raman, Adam B. Weiner

## Abstract

**Background:** The incremental value of PSMA PET/CT over multiparametric MRI (mpMRI) for selecting patients for prostate cancer focal therapy is undefined. Unlike lesion-detection studies, we evaluated whether PSMA PET/CT, mpMRI, and their combination correctly determine focal therapy eligibility when validated against whole-mount histopathology (WMHP).

**Methods:** We retrospectively analyzed patients undergoing radical prostatectomy (RP) at UCLA (2017-2024) with preoperative PSMA PET/CT and mpMRI, biopsy grade group (GG) ≤3, unilateral disease, and prostate-specific antigen <20 ng/mL. Two blinded readers per modality assessed extraprostatic extension (EPE), seminal vesicle invasion (SVI), bilateral disease, and multifocality; a third adjudicated discordances. Eligibility required absence of all four. On WMHP, ineligibility was EPE, SVI, multifocal or bilateral disease with a contralateral or secondary GG≥2 nodule, or any GG>3 focus at RP. Modalities were compared by McNemar tests.

**Results:** Of 112 patients, 77 (69%) were ineligible on WMHP, including 19 (17%) with a GG>3 focus at RP. Sensitivity and specificity were 53% and 54% for PET and 70% and 49% for mpMRI. Combined imaging improved sensitivity to 83% (*P*<0.001 vs PET; *P*=0.002 vs MRI) but reduced specificity to 26%. Per 100 patients at this prevalence, this would correctly identify approximately 9 additional ineligible patients while falsely excluding approximately 7 true candidates. mpMRI was more sensitive than PET for EPE (77% vs 32%, *P*<0.001), whereas PET was more specific (90% vs 58%, *P*<0.001). Combined imaging improved detection of multifocal (44% vs 23%, *P*=0.001) and bilateral disease (45% vs 23%, *P*=0.002) versus MRI alone.

**Conclusions:** Adding PSMA PET/CT to mpMRI significantly improves sensitivity for exclusionary features, reflecting complementary molecular and anatomic information. However, combined imaging still misses a substantial proportion of adverse pathology, so imaging alone should not determine eligibility.

Graphical Abstract

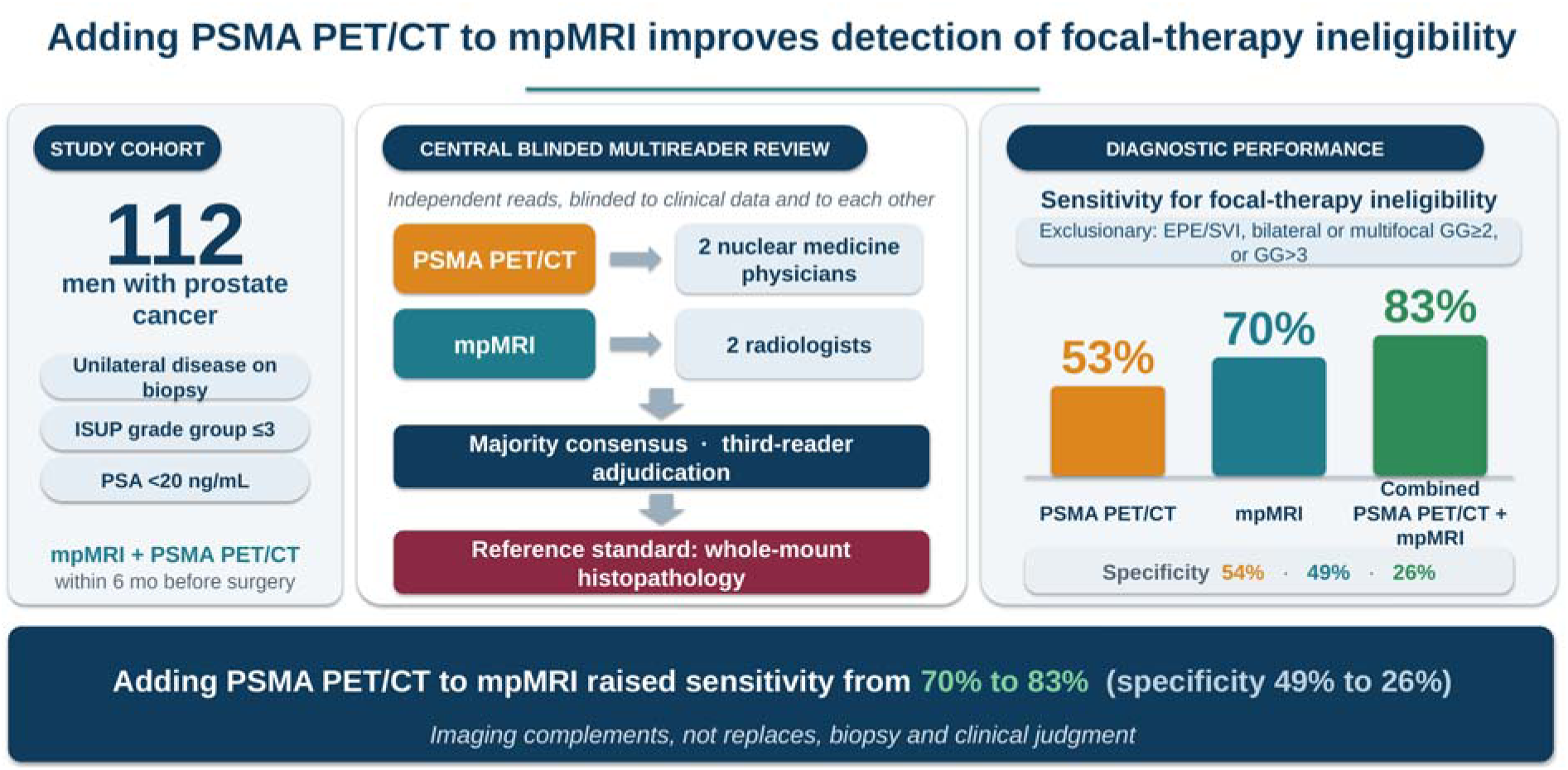

## Introduction

Focal therapy of prostate cancer is of increasing interest because it may provide durable cancer control while preserving quality of life relative to whole-gland treatment.^1–3^ However, the success of focal therapy depends on accurate pretreatment characterization of intraprostatic disease, including tumor focality, laterality, and the absence of extraprostatic extension (EPE) or seminal vesicle invasion (SVI).^4,5^ Although in-field recurrences occur, inadequate disease mapping is a recognized contributor to oncologic failure after focal therapy.^5^

Multiparametric MRI (mpMRI) is currently the cornerstone of focal therapy planning.^5,6^ Although mpMRI improves detection of clinically significant lesions, it may underestimate disease extent, particularly tumor volume and multifocality.^7^ Johnson et al. found that 48% of men deemed hemiablation candidates by mpMRI and fusion biopsy harbored contralateral significant or ipsilateral high-grade disease on whole-mount histopathology (WMHP).^8^ Prostate-specific membrane antigen positron emission tomography/computed tomography (PSMA PET/CT) has emerged as a sensitive modality for prostate cancer detection and staging^9,10^, with recent data demonstrating complementary detection patterns with mpMRI.^11^ There is growing interest in its role for primary disease assessment and focal therapy planning.^12^

Whether molecular imaging with PSMA PET/CT adds to mpMRI for this composite, eligibility-level task, validated against definitive surgical pathology, remains undefined. We hypothesized that PSMA PET/CT and mpMRI provide complementary information and that their combination improves detection of exclusionary disease features. We therefore evaluated the diagnostic performance of PSMA PET/CT, mpMRI, and their combination for determining focal therapy eligibility using WMHP as the reference standard in a surgically validated cohort.

## Materials and Methods

### Patient Selection and Study Design

We conducted a single-center, retrospective study of patients who underwent mpMRI and PSMA PET within 6 mo prior to radical prostatectomy at UCLA between January 2017 and July 2024. Patients were identified from a prospectively maintained institutional database (**Supplemental Fig. 1**). Of 420 patients screened, 130 (31%) met inclusion criteria: International Society of Urological Pathology grade group (GG) ≤3 and unilateral disease on biopsy and prostate-specific antigen (PSA) <20 ng/mL. Of these, 18 (14%) were subsequently excluded for incomplete imaging or histopathologic data (n = 10), other missing data (n = 7), or unassignable Gleason grade due to neoadjuvant hormonal therapy (n = 1), yielding a final cohort of 112 patients. The study was approved by the UCLA institutional review board (10-1863), and the need for written informed consent was waived given the retrospective design. This report follows the Standards for Reporting of Diagnostic Accuracy Studies (STARD) guidelines (**Appendix**). Prostate biopsy at diagnosis followed institutional practice: MRI-targeted sampling of any PI-RADS ≥3 lesion together with systematic sampling, or systematic sampling alone when no lesion was identified. Where the approach was recorded (55 of 112), this was MRI–ultrasound fusion (n = 35), transrectal ultrasound–guided systematic biopsy (n = 16), or in-bore MRI-guided biopsy (n = 4). Core counts were incompletely captured (median systematic cores, 11 [IQR, 10–12; n = 65]; median targeted cores, 3 [IQR, 3–5; n = 47]). The transperineal versus transrectal approach was not recorded, and both were in use at our institution during the study period. Because focal therapy candidacy rests on biopsy-based disease mapping, the extent of systematic and contralateral sampling influences how many patients are judged eligible before surgery.^13^ PSMA PET/CT was obtained as part of routine clinical practice rather than under a study protocol, typically in men with unfavorable intermediate– or high-risk disease. This referral pattern is itself a source of selection bias and is revisited in the limitations.

### PSMA PET and mpMRI Acquisition

PSMA PET/CT was performed using [^68^Ga]Ga-PSMA-11 or [^18^F]DCFPyL on Siemens Biograph mCT or Vision PET/CT scanners (Siemens Healthineers), as previously described for [^68^Ga]Ga-PSMA-11.^14^ mpMRI was performed on 3T MRI systems (Siemens Healthineers) with T2-weighted imaging, diffusion-weighted imaging, and dynamic contrast-enhanced MRI, interpreted per the Prostate Imaging Reporting and Data System (PI-RADS) v2.1.^15^

### Imaging Interpretation

Two board-certified nuclear medicine physicians (A.H., S.B.) independently reviewed PSMA PET scans and two board-certified radiologists (A.C., T.S.) independently reviewed mpMRI studies, blinded to clinical data and to each other (PSA was provided). Readers assessed EPE, SVI, bilaterality, and multifocality (two or more spatially separate foci). PET interpretation used expert visual assessment; PRIMARY ≥3 and PI-RADS ≥3 lesions were positive for lesion detection (**Figure 1**). Eligibility required absence of all four. Discordances were adjudicated by a third reader (I.S. and S.R. for PET and MRI, respectively). The combined PET + MRI result was defined a priori as imaging-ineligible if either modality identified any exclusionary feature. An independent dual board-certified reader (K.K.) performed an integrated read (**Supplemental Methods**).

**Figure 1.**
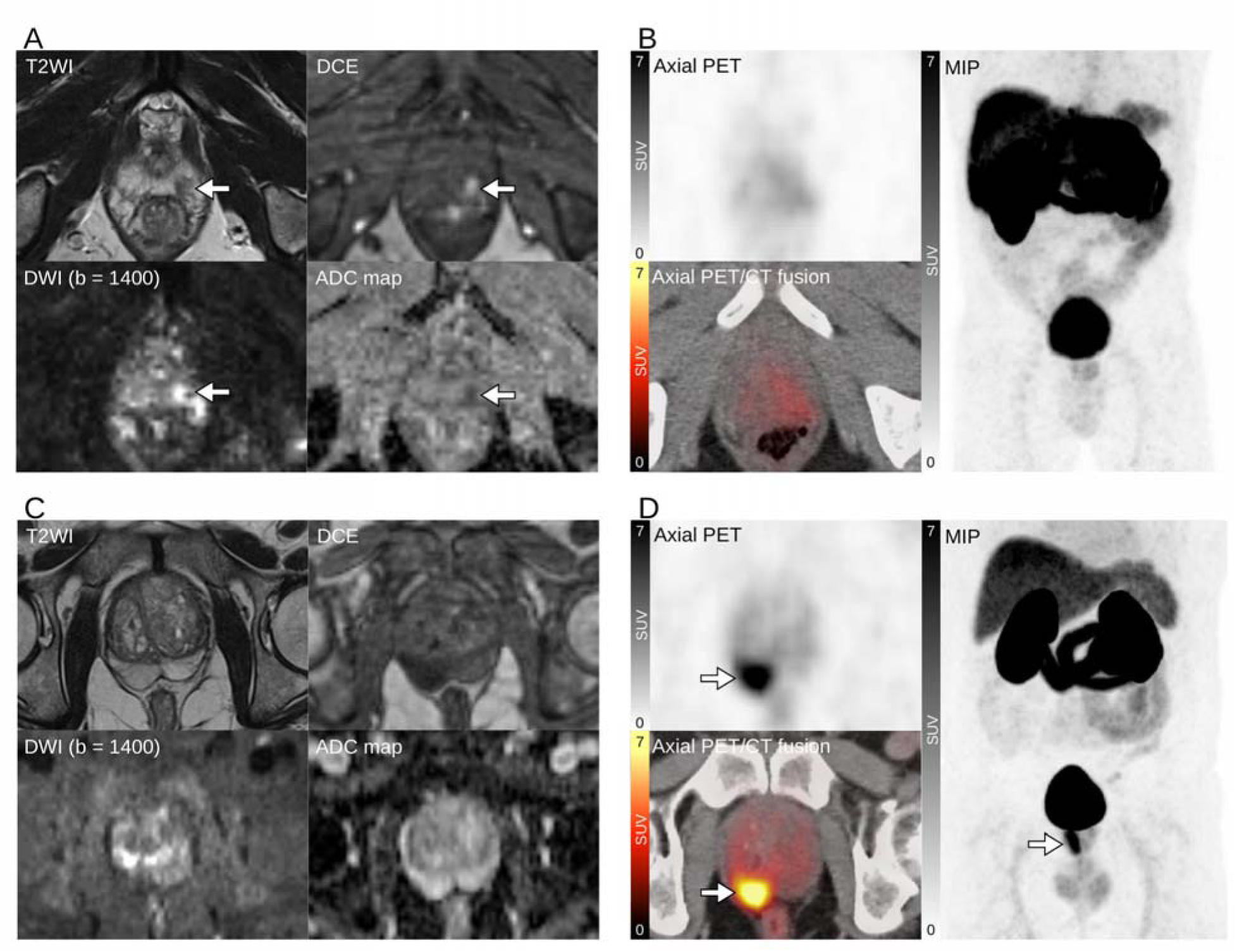
Complementary detection patterns of multiparametric MRI (mpMRI) and PSMA PET/CT in two representative patients. (A and B) Case 1, a patient with an MRI-positive but PSMA PET/CT-negative lesion. (A) mpMRI demonstrates a lesion (arrows) in the left posterolateral peripheral zone at the apex, with low signal intensity on T2WI, diffusion restriction on DWI (b = 1400) and the ADC map, and early enhancement on DCE imaging, consistent with PI-RADS category 4. (B) Corresponding PSMA PET/CT (axial PET, axial PET/CT fusion, and MIP) shows no abnormal tracer uptake in the prostate, corresponding to a PRIMARY score of 1. Whole-mount histopathology confirmed Gleason score 3+4 = 7 (grade group 2) cancer at the site of the MRI lesion, unifocal and organ-confined, with no seminal vesicle invasion. (C and D) Case 2, a patient with an MRI-negative but PSMA PET/CT-positive lesion. (C) mpMRI demonstrates no findings suspicious for clinically significant cancer (PI-RADS score ≤2). (D) PSMA PET/CT demonstrates intense focal tracer uptake (arrows; SUVmax 10.4) in the right posterior peripheral zone extending from the base to the apex, corresponding to a PRIMARY score of 4. Whole-mount histopathology confirmed Gleason score 4+3 = 7 (grade group 3) adenocarcinoma with extensive cribriform architecture and intraductal carcinoma, with extraprostatic extension at the site of tracer uptake. ADC = apparent diffusion coefficient; DCE = dynamic contrast-enhanced; DWI = diffusion-weighted imaging; MIP = maximum-intensity projection; mpMRI = multiparametric MRI; PI-RADS = Prostate Imaging Reporting and Data System; PSMA = prostate-specific membrane antigen; SUVmax = maximum standardized uptake value; T2WI = T2-weighted imaging.

### Histopathologic Reference Standard

WMHP from prostatectomy specimens served as the reference standard, reviewed by a genitourinary pathologist (A.S.). Ineligibility was defined as the presence of ≥1 of the following: EPE, SVI, multifocal or bilateral disease with a contralateral or secondary GG≥2 nodule, or any GG>3 focus at RP. GG4-5 patterns confer biologic aggressiveness, and focal therapy is generally offered for them only with caution.^5^ Patients with only contralateral GG1 disease (and no high-grade or extraprostatic features) were eligible, consistent with current surveillance practice for GG1.^16,17^ A stricter sensitivity analysis retained any (including GG1) bilateral or multifocal disease as exclusionary.

### Statistical Analysis

The primary outcome was sensitivity for detecting ineligibility for focal therapy. Specificity, positive predictive value (PPV), and negative predictive value (NPV) were secondary outcomes. Ninety-five percent confidence intervals (CIs) for proportions were calculated using the exact (Clopper-Pearson) method. Paired comparisons of sensitivity and specificity between modalities used McNemar tests. Exploratory subgroup analyses were restricted to patients with biopsy grade group 1-2 disease, the lower-risk group most often considered for focal therapy, and, paralleling Johnson et al.^8^, to patients eligible by mpMRI alone. Inter-reader agreement used Cohen’s κ (**Supplemental Methods**). All P values were two-sided, and P<0.05 was considered significant. Analyses used R 4.6.0.

## Results

### Patient and Tumor Characteristics

Of the 112 patients included, median age was 65 yr (interquartile range [IQR], 59-69) and median PSA was 6.7 ng/mL (IQR, 4.8-9.4; **Table 1**). The median interval between mpMRI and PSMA PET was 66 d (IQR, 38-89); the median interval from the later imaging study to radical prostatectomy was 56 d (IQR, 34-82). On WMHP, 77 patients (69%) were ineligible under the primary definition, including 19 (17%) with a GG>3 focus (15 GG5, 4 GG4); feature-level frequencies are given in **Table 1**.

**Table 1.** Patient Characteristics (N = 112)

| Characteristic | Value |
| --- | --- |
| Age, yr, median (IQR) | 65 (59-69) |
| PSA, ng/mL, median (IQR) | 6.7 (4.8-9.4) |
| Biopsy GG, n (%) |  |
| GG1 (3+3) | 5 (4) |
| GG2 (3+4) | 30 (27) |
| GG3 (4+3) | 77 (69) |
| NCCN risk group, n (%) |  |
| Low | 5 (4) |
| Favorable intermediate | 29 (26) |
| Unfavorable intermediate | 78 (70) |
| mpMRI-PET interval, d, median (IQR) | 66 (38-89) |
| Imaging-to-prostatectomy interval, d, median (IQR) | 56 (34-82) |
| WMHP findings, n (%) |  |
| EPE | 53 (47) |
| SVI | 18 (16) |
| Bilateral disease | 47 (42) |
| Multifocal disease | 52 (46) |
| Any GG>3 focus at RP | 19 (17) |
| Ineligible for focal therapy | 77 (69) |

EPE = extraprostatic extension; GG = grade group; IQR = interquartile range; mpMRI = multiparametric MRI; NCCN
= National Comprehensive Cancer Network; PSA = prostate-specific antigen; RP = radical prostatectomy; SVI = seminal vesicle invasion; WMHP = whole-mount histopathology.

### Focal Therapy Eligibility by Imaging

For the composite eligibility endpoint, PSMA PET had a sensitivity of 53% and a specificity of 54% (PPV 72%, NPV 35%), whereas mpMRI had a sensitivity of 70% and a specificity of 49% (PPV 75%, NPV 42%) (**Table 2**; **Figure 2**). mpMRI sensitivity was higher than that of PET (P=0.035). Combining both modalities improved sensitivity to 83% (P<0.001 vs PET; P=0.002 vs MRI), though specificity decreased to 26% (P=0.002 vs PET; P=0.008 vs MRI). The single-reader integrated interpretation was broadly concordant (**Supplemental Table 1**); reader-level score distributions are given in **Supplemental Table 2**. Representative discordant cases are shown in **Figure 1**.

**Figure 2.**
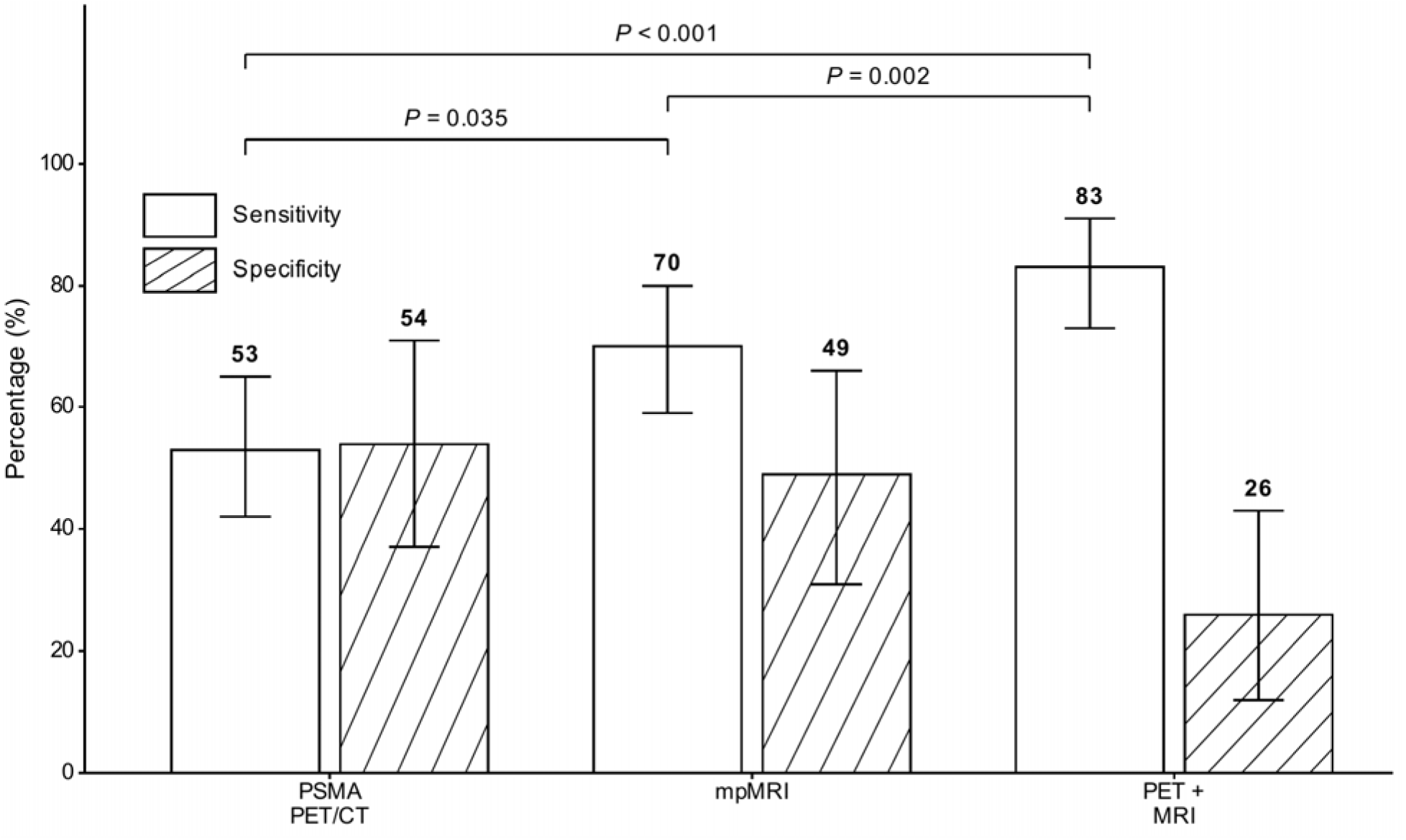
Diagnostic performance for focal therapy eligibility by imaging modality. Bars show point estimates with error caps spanning the 95% exact binomial (Clopper-Pearson) confidence interval. The PET + MRI column reflects a rule-based combination (positive if either modality was positive by majority read). Comparisons use the McNemar test.

**Table 2.**
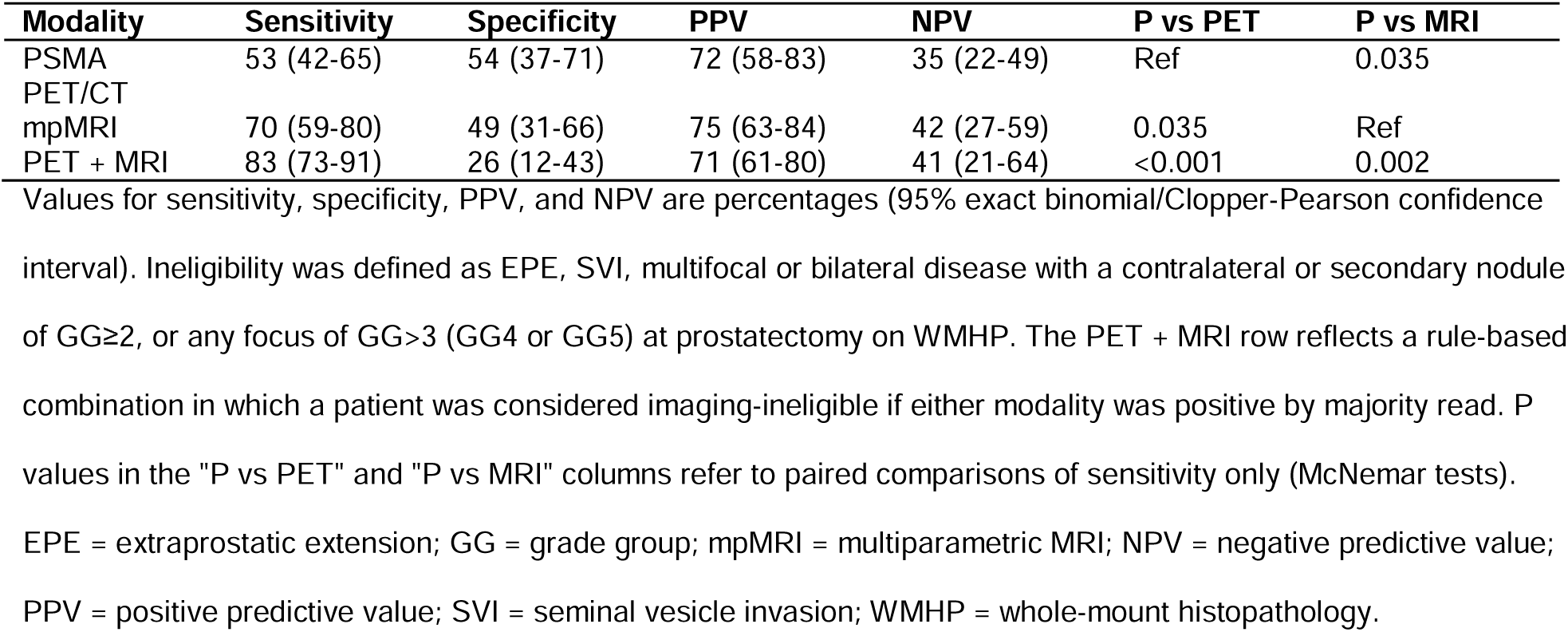
Diagnostic Performance for Focal Therapy Eligibility (Composite Endpoint)

### Detection of Individual Pathologic Features

For EPE, mpMRI had significantly higher sensitivity than PET (77% vs 32%, *P*<0.001), whereas PET had higher specificity (90% vs 58%, *P*<0.001) (**Table 3**; **Figure 3**). For SVI, both modalities performed comparably, with high specificity (96-99%) and moderate sensitivity (PET 39%, MRI 44%; *P*=1.00). For multifocal and bilateral disease, PET was more sensitive and MRI more specific, though not significantly so. Adding PET to MRI improved sensitivity for multifocal disease (44% vs 23%, *P*=0.001) and bilateral disease (45% vs 23%, *P*=0.002) compared with MRI alone. For the high-grade criterion (17% prevalence), imaging-based ineligibility captured 58%, 74%, and 79% of cases for PET, mpMRI, and PET + mpMRI. Inter-reader agreement was higher for mpMRI (κ 0.48-0.60) than PSMA PET (κ 0.02-0.48), particularly for EPE (**Supplemental Table 3**).

**Figure 3.**
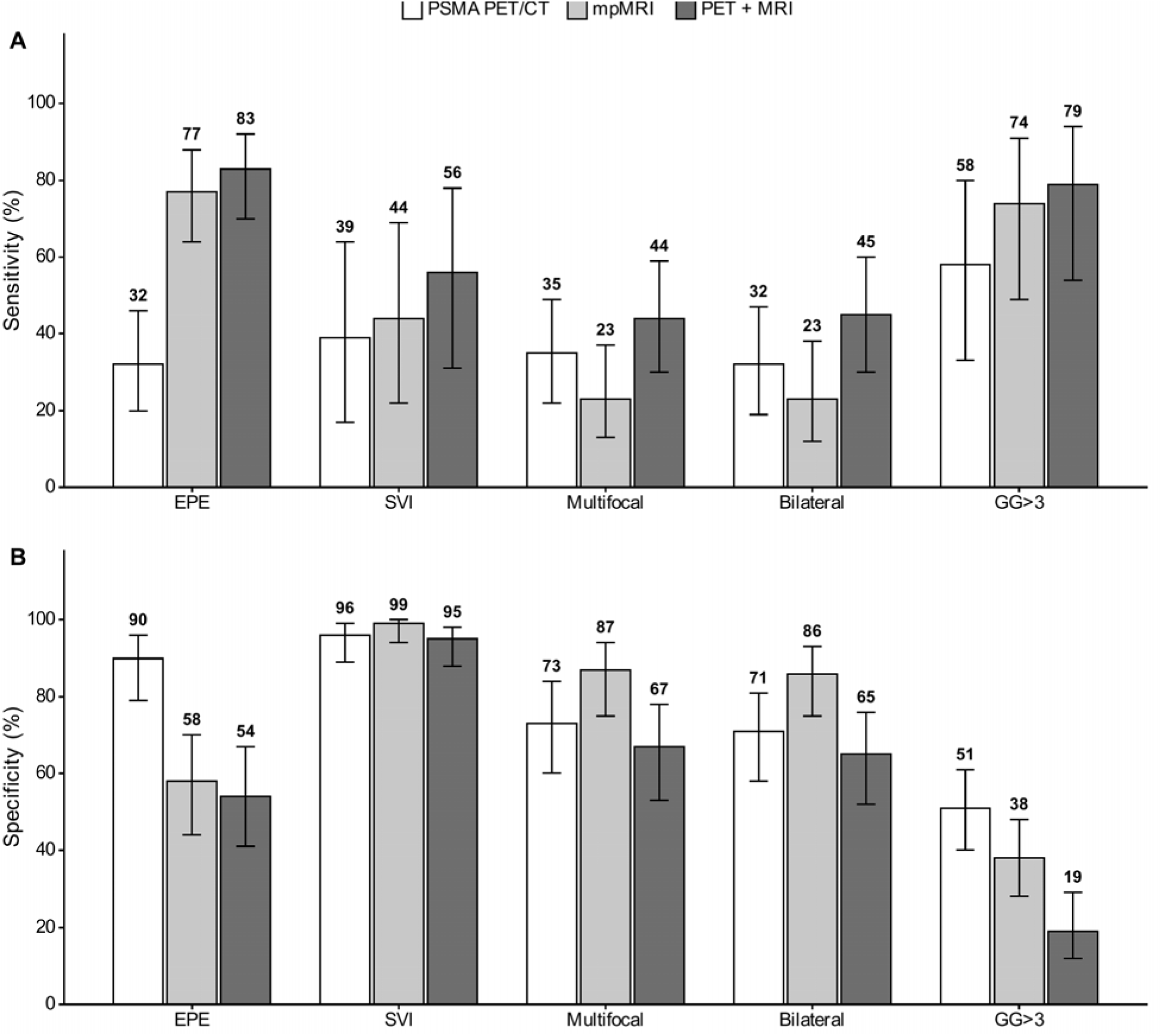
Sensitivity (A) and specificity (B) of imaging modalities across individual pathologic features. Bars show point estimates with error caps spanning the 95% exact binomial (Clopper-Pearson) confidence interval. The PET + MRI bars reflect a rule-based combination (positive if either modality was positive by majority read). For EPE, SVI, multifocal, and bilateral disease, predictions reflect the corresponding per-feature imaging call. For GG>3 (which imaging did not directly assess), bars reflect each modality’s composite ineligibility call. PSMA PET/CT and mpMRI demonstrated complementary diagnostic profiles. Pairwise comparisons by McNemar test (see Table 3). EPE = extraprostatic extension; GG = grade group; mpMRI = multiparametric MRI; SVI = seminal vesicle invasion.

**Table 3.** Diagnostic Performance for Individual Pathologic Features.

| Feature | Modality | Sens | Spec | PPV | NPV | P (Sens) | P (Spec) |
| --- | --- | --- | --- | --- | --- | --- | --- |
| EPE | PET | 32 (20-46) | 90 (79-96) | 74 (52-90) | 60 (49-70) | Ref | Ref |
|  | MRI | 77 (64-88) | 58 (44-70) | 62 (49-74) | 74 (59-86) | <0.001 | <0.001 |
|  | PET+MRI | 83 (70-92) | 54 (41-67) | 62 (50-73) | 78 (62-89) | - | - |
| SVI | PET | 39 (17-64) | 96 (89-99) | 64 (31-89) | 89 (81-94) | Ref | Ref |
|  | MRI | 44 (22-69) | 99 (94-100) | 89 (52-100) | 90 (83-95) | 1.00 | 1.00 |
|  | PET+MRI | 56 (31-78) | 95 (88-98) | 67 (38-88) | 92 (84-96) | - | - |
| Multifocal | PET | 35 (22-49) | 73 (60-84) | 53 (35-70) | 56 (45-68) | Ref | Ref |
|  | MRI | 23 (13-37) | 87 (75-94) | 60 (36-81) | 57 (46-67) | 0.21 | 0.077 |
|  | PET+MRI | 44 (30-59) | 67 (53-78) | 49 (34-64) | 55 (43-68) | - | - |
| Bilateral | PET | 32 (19-47) | 71 (58-81) | 44 (27-62) | 59 (47-70) | Ref | Ref |
|  | MRI | 23 (12-38) | 86 (75-93) | 55 (32-77) | 61 (50-71) | 0.45 | 0.031 |
|  | PET+MRI | 45 (30-60) | 65 (52-76) | 48 (32-63) | 62 (49-73) | - | - |
| GG>3 | PET | 58 (33-80) | 51 (40-61) | 19 (9-20,22-32) | 85 (73-94) | Ref | Ref |
|  | MRI | 74 (49-91) | 38 (28-48) | 19 (10-20,22-29,32) | 88 (73-96) | 0.375 | 0.104 |
|  | PET+MRI | 79 (54-94) | 19 (11-20,22-28,32) | 17 (9-20,22-26) | 82 (60-95) | - | - |

Values for Sens, Spec, PPV, and NPV are percentages (95% exact binomial/Clopper-Pearson confidence interval).
The PET+MRI row reflects a rule-based combination in which a patient was considered positive for a given feature if either modality was positive by majority read. For EPE, SVI, multifocality, and bilaterality, predictions reflect the corresponding per-feature imaging call. Because grade was not directly assessed on imaging, the GG>3 row uses each modality's composite ineligibility call as the prediction, capturing high-grade patients via coincident adverse imaging features. P values from McNemar tests compare paired sensitivity and specificity between PSMA PET/CT and mpMRI only; PET + MRI values are shown for reference but were not used as the comparator. EPE = extraprostatic extension; GG = grade group; mpMRI = multiparametric MRI; NPV = negative predictive value; PPV = positive predictive value; Sens = sensitivity; Spec = specificity; SVI = seminal vesicle invasion.

### Sensitivity Analysis

Under a stricter definition in which any bilateral or multifocal disease was exclusionary, 90 of 112 patients (80%) were ineligible, with modestly lower sensitivity across modalities (**Supplemental Table 4**).

### Lower-Risk Subgroup Analysis

Among the 35 patients with biopsy grade group 1-2 disease, 22 (63%) were ineligible by WMHP; combined imaging reached 86% sensitivity and 62% NPV (**Supplemental Table 5**).

### Secondary Analysis: mpMRI-Eligible Subgroup

To parallel the design of Johnson et al.^8^, we performed a secondary analysis restricted to the 33 patients (29%) deemed eligible for focal therapy by mpMRI alone. On WMHP, 20 of these 33 patients (61%) were ineligible, harboring at least one exclusionary feature. Adding PSMA PET reclassified 15 of 33 mpMRI-eligible patients (45%) as ineligible.

## Discussion

Reported as early as 2006^18^, focal therapy for prostate cancer is now increasingly used, although both the National Comprehensive Cancer Network and the European Association of Urology do not recommend it as initial treatment outside clinical trials or prospective registries.^16,17^ However, treatment failure remains a major concern, with in-field failure of 24-35% within 18 months in a contemporary series^19^ and wide variation across modalities in systematic review.^20^ In this surgically validated cohort, we found that 69% of patients meeting contemporary criteria harbored exclusionary features on WMHP, including 17% with high-grade (GG>3) disease at prostatectomy.

Our findings confirm that mpMRI and PSMA PET have distinct and complementary diagnostic profiles. mpMRI demonstrated higher sensitivity for EPE, consistent with prior surgical correlation studies^21^, but showed limited sensitivity for multifocal and bilateral disease. This echoes Johnson et al., in whose series nearly half of apparent hemiablation candidates had disqualifying disease on WMHP.^8^ PSMA PET showed numerically higher sensitivity for multifocal and bilateral disease, though these differences were not statistically significant, and mpMRI was significantly more specific. These findings extend PRIMARY, which established complementary patient-level detection^22^, and PRIMARY2, which addressed biopsy avoidance^23^, to the distinct task of whole-mount-validated eligibility determination.

At RP, GG>3 was noted in 19 patients (17%). Most (18 of 19) were already ineligible via other criteria. The remaining patient had a unifocal unilateral GG5 nodule without EPE or SVI and would otherwise have appeared a candidate. Because neither modality directly grades tumor foci, both capture this criterion only via coincident adverse imaging features, which drove the modest decreases in composite sensitivity versus a grade-agnostic endpoint. The mpMRI EPE sensitivity of 77% exceeds some prior surgical series, likely reflecting enrichment for bulky macroscopic EPE among surgical candidates; microscopic EPE remains poorly detected.^21^

Combining PSMA PET and mpMRI significantly improved sensitivity (83% vs 53% for PET and 70% for MRI), driven primarily by better detection of multifocal and bilateral disease when adding PET to MRI. The trade-off was reduced specificity. At the ineligibility prevalence observed here (69%), moving from mpMRI alone to combined imaging would, per 100 patients evaluated, correctly identify approximately 9 additional patients with exclusionary pathology while falsely excluding approximately 7 additional true candidates. Because a falsely excluded patient is redirected to whole-gland therapy or further workup whereas a falsely included patient is treated with undetected residual disease, we regard this exchange as clinically acceptable, though the balance would shift in a lower-prevalence population.

Inadequate pretreatment characterization is one contributor to focal therapy failure, alongside technical and biologic factors.^5,24^ Applying the framework of Johnson et al.^8^ to our cohort, 61% of mpMRI-eligible patients had exclusionary features on WMHP. PSMA PET correctly reclassified 8 of these but incorrectly excluded 7 true candidates, and 67% of patients eligible by both modalities remained ineligible on WMHP. These data argue against imaging-only eligibility determination and support investigation of improved imaging modalities and better biopsy techniques.^25^ Our findings are consistent with integrated imaging-biopsy models. Geboers et al. reported that adding PSMA PET to mpMRI and systematic biopsy improved discrimination for clinically significant cancer (area under the curve, 0.79 to 0.84) and correctly excluded 26 of 46 patients unsuitable for hemiablation with a 3% false-exclusion rate.^12^ Comparable complementarity has since been described in focal therapy candidate selection.^26,27^ The limits of biopsy-based mapping are likewise well documented, from template mapping^13^ to the PROMIS and PICTURE analyses of MRI-undetected cancer.^28^

Inter-reader agreement differed substantially between modalities. mpMRI readers reached moderate agreement (Cohen’s κ 0.48-0.60), whereas PSMA PET readers achieved only fair-to-chance agreement (κ as low as 0.02 for EPE and 0.04 for composite eligibility). This is lower than the reproducibility reported for the PRIMARY score, for which Emmett et al. found substantial interreader agreement across six readers (κ 0.70 for the binary score vs 0.58 for PI-RADS).^29^ The discrepancy is instructive rather than contradictory: PRIMARY was developed for intraprostatic lesion detection, whereas our readers judged extraprostatic extension, seminal vesicle invasion, and laterality, tasks for which no validated PSMA PET reporting framework yet exists. Standardized PSMA PET reporting and dedicated reader training will likely be required before molecular imaging can be relied upon for eligibility decisions.

In the lower-risk subgroup, 63% still had exclusionary features on WMHP, indicating that the limits of biopsy-based selection extend to lower-risk patients; wide confidence intervals warrant prospective validation.

The retrospective, single-center design limits generalizability. Restricting the cohort to men who had a PSMA PET before prostatectomy selects for higher-risk patients who elected surgery; consequently 69% were ineligible on WMHP, well above the rate expected in a typical focal therapy evaluation. PPV and NPV are influenced by prevalence and likely do not generalize to broader focal therapy populations. In addition, because imaging spanned 2017 to 2024, earlier-generation acquisition and evolving PSMA reporting criteria may have attenuated measured performance relative to contemporary protocols.

## Conclusions

Neither PSMA PET/CT nor mpMRI alone reliably identified focal therapy candidates when compared with WMHP. Combining both modalities significantly improved sensitivity, driven by complementary detection of pathologic features. These findings support a dual-modality imaging strategy for focal therapy patient selection; however, a considerable proportion of patients remain inaccurately characterized. Prospective studies integrating imaging with biopsy data and emerging biomarkers are needed.

## Data Availability

The datasets generated and analyzed during the current study are not publicly available owing to institutional privacy restrictions but are available from the corresponding author on reasonable request.

## Acknowledgements

Data and research support were provided by the Integrated Diagnostics (IDx) Shared Resource, Department of Radiological Sciences, David Geffen School of Medicine at UCLA. Generative artificial intelligence (Claude; Anthropic) was used to assist with manuscript formatting and language editing; the authors reviewed all output and take full responsibility for the content of the publication.

## Key Points

### Question

Can PSMA PET/CT, multiparametric MRI, or their combination accurately determine focal therapy eligibility in prostate cancer when validated against whole-mount histopathology? **Pertinent Findings:** Among 112 men undergoing radical prostatectomy with preoperative PSMA PET/CT and multiparametric MRI, 69% had exclusionary disease on whole-mount histopathology. Combining modalities raised sensitivity for ineligibility to 83%, above multiparametric MRI (70%) or PSMA PET/CT (53%) alone, although specificity fell to 26%.

### Implications for Patient Care

Dual-modality PSMA PET/CT and multiparametric MRI improves identification of patients unsuitable for focal therapy, but imaging alone remains insufficient and should be combined with biopsy data and clinical judgment.

## Supplemental material

Supplemental Figure 1; Supplemental Tables 1-5; Supplemental Methods and Results; STARD checklist (Appendix)

## Prior presentation

Presented in part at the Society of Abdominal Radiology Annual Meeting 2025.

## References

1. Valerio M, Cerantola Y, Eggener SE. New and established technology in focal ablation of the prostate: a systematic review. Eur Urol. 2017;71:17–34.

2. Azzouzi AR, Vincendeau S, Barret E. Padeliporfin vascular-targeted photodynamic therapy versus active surveillance in men with low-risk prostate cancer (CLIN1001 PCM301): an open-label, phase 3, randomised controlled trial. Lancet Oncol. 2017;18:181–191.

3. Reddy D, Peters M, Shah TT. Cancer control outcomes following focal therapy using high-intensity focused ultrasound in 1379 men with nonmetastatic prostate cancer: a multi-institute 15-year experience. Eur Urol. 2022;81:407–413.

4. Oishi M, Gill IS, Tafuri A. Hemigland cryoablation of localized low, intermediate and high risk prostate cancer: oncologic and functional outcomes at 5 years. J Urol. 2019;202:1188–1198.

5. Ghoreifi A, Gomella L, Hu JC. Identifying the best candidate for focal therapy: a comprehensive review. Prostate Cancer Prostatic Dis. 2025;28:684–692.

6. Kasivisvanathan V, Rannikko AS, Borghi M. MRI-targeted or standard biopsy for prostate-cancer diagnosis. N Engl J Med. 2018;378:1767–1777.

7. Le JD, Tan N, Shkolyar E. Multifocality and prostate cancer detection by multiparametric magnetic resonance imaging: correlation with whole-mount histopathology. Eur Urol. 2015;67:569–576.

8. Johnson DC, Yang JJ, Kwan L. Do contemporary imaging and biopsy techniques reliably identify unilateral prostate cancer? Implications for hemiablation patient selection. Cancer. 2019;125:2955–2964.

9. Hofman MS, Lawrentschuk N, Francis RJ, et al. Prostate-specific membrane antigen PET-CT in patients with high-risk prostate cancer before curative-intent surgery or radiotherapy (proPSMA): a prospective, randomised, multicentre study. Lancet Lond Engl. 2020;395(10231):1208–1216. doi:10.1016/S0140-6736(20)30314-7

10. Sonni I, Eiber M, Fendler WP. Impact of 68Ga-PSMA-11 PET/CT on staging and management of prostate cancer patients in various clinical settings: a prospective single-center study. J Nucl Med. 2020;61:1153–1160.

11. Sonni I, Weiner AB, Doddipalli S, et al. Clinical, Pathologic, and Imaging Variables Associated with Prostate Cancer Detection by PSMA PET/CT and Multiparametric MRI. J Nucl Med Off Publ Soc Nucl Med. 2024;65(12):1923–1931. doi:10.2967/jnumed.124.268443

12. Geboers B, Meijer D, Counter W. Prostate-specific membrane antigen positron emission tomography in addition to multiparametric magnetic resonance imaging and biopsies to select prostate cancer patients for focal therapy. BJU Int. 2024;133(suppl 4):14–22.

13. Valerio M, Anele C, Freeman A. Identifying the index lesion with template prostate mapping biopsies. J Urol. 2015;193:1185–1190.

14. Calais J, Ceci F, Eiber M. 18F-fluciclovine PET-CT and 68Ga-PSMA-11 PET-CT in patients with early biochemical recurrence after prostatectomy: a prospective, single-centre, single-arm, comparative imaging trial. Lancet Oncol. 2019;20:1286–1294.

15. Turkbey B, Rosenkrantz AB, Haider MA. Prostate Imaging Reporting and Data System version 2.1: 2019 update of Prostate Imaging Reporting and Data System version 2. Eur Urol. 2019;76:340–351.

16. Spratt DE, Srinivas S, Adra N, et al. NCCN Guidelines® Insights: Prostate Cancer, Version 5.2026: Featured Updates to the NCCN Guidelines. J Natl Compr Canc Netw. 2026;24(5):140–149. doi:10.6004/jnccn.2026.0023

17. Cornford P, Van Den Bergh RCN, Briers E, et al. EAU-EANM-ESTRO-ESUR-ISUP-SIOG Guidelines on Prostate Cancer—2024 Update. Part I: Screening, Diagnosis, and Local Treatment with Curative Intent. Eur Urol. 2024;86(2):148–163. doi:10.1016/j.eururo.2024.03.027

18. Bahn DK, Silverman P, F L Sr, Badalament R, Bahn ED, Rewcastle JC. Focal prostate cryoablation: initial results show cancer control and potency preservation. J Endourol. 2006;20:688–692.

19. Aker MN, Brisbane WG, Kwan L. Cryotherapy for partial gland ablation of prostate cancer: oncologic and safety outcomes. Cancer Med. 2023;12:9351–9362.

20. Hopstaken JS, Bomers JGR, Sedelaar MJP, Valerio M, Fütterer JJ, Rovers MM. An updated systematic review on focal therapy in localized prostate cancer: what has changed over the past 5 years? Eur Urol. 2022;81:5–33.

21. M R, EHJ H, JA W, JO B, MM R. Accuracy of magnetic resonance imaging for local staging of prostate cancer: a diagnostic meta-analysis. Eur Urol. 2016;70:233–245.

22. Emmett L, Buteau J, Papa N. The additive diagnostic value of prostate-specific membrane antigen positron emission tomography computed tomography to multiparametric magnetic resonance imaging triage in the diagnosis of prostate cancer (PRIMARY): a prospective multicentre study. Eur Urol. 2021;80:682–689.

23. Buteau JP, Moon D, Fahey MT. Effect of [68Ga]Ga-PSMA-11 PET-CT in the diagnosis of prostate cancer in men with equivocal or clinically high-risk non-suspicious findings on multiparametric MRI (PRIMARY2): a multicentre, non-inferiority, phase 3, randomised controlled trial. Lancet Oncol. 2026;27:839–848.

24. Shah TT, Peters M, Eldred-Evans D. Early-medium-term outcomes of primary focal cryotherapy to treat nonmetastatic clinically significant prostate cancer from a prospective multicentre registry. Eur Urol. 2019;76:98–105.

25. Weiner AB, Proudfoot JA, Aker M. Genomic biomarker for prostate cancer focal therapy: post hoc assessment of a phase II clinical trial. JCO Precis Oncol. 2025;9:e2500535.

26. Lim KY, Asghari-Jafarabadi M, Kalapara AA, Smith J, Frydenberg M, Ranasinghe W. The addition of PSMA PET in selecting candidates for prostate cancer focal therapy. BJUI Compass. 2026;7:e70264.

27. Stabile A, Brancaccio M, Scilipoti P. Patient selection for prostate cancer focal therapy guided by mpMRI-PSMA PET concordance and pathology review. BJU Int August. 2026;11.

28. Light A, Peters M, Gopalakrishnan A. Concomitant MRI-detected and MRI-undetected prostate cancer with implications for biopsy strategies and focal therapy planning: an analysis of the PROMIS and PICTURE trials. Eur Urol Oncol June. 2026;16.

29. Emmett L, Papa N, Counter W. Reproducibility and accuracy of the PRIMARY score on PSMA PET and of PI-RADS on multiparametric MRI for prostate cancer diagnosis within a real-world database. J Nucl Med. 2024;65:94–99.

