## Supplemental Data 1 for "Prostate-Specific Membrane Antigen PET/CT Complements MRI for Focal Therapy Selection in Prostate Cancer: A Whole-Mount Histopathologic Study"

**Description of Supplementary Material**

File format: Microsoft Word (.docx). This single file contains all supplementary items listed below; each is self-contained and directly relevant to the conclusions of the main article.

Supplemental Figure 1 (≤50 words): Patient selection flowchart showing screening of 420 patients to the final cohort of 112, with reasons for exclusion.

Supplemental Table 1 (≤50 words): Diagnostic performance of the integrated PET + MRI read by a single dual-board-certified reader, versus the rule-based combination and each single modality.

Supplemental Table 2 (≤50 words): Distribution of PRIMARY and PI-RADS scores by reader (n=112).

Supplemental Table 3 (≤50 words): Inter-reader agreement (Cohen’s κ) between PET and MRI reader pairs for each exclusionary feature and the composite endpoint.

Supplemental Table 4 (≤50 words): Sensitivity analysis under a stricter eligibility definition retaining any bilateral or multifocal disease as exclusionary.

Supplemental Table 5 (≤50 words): Diagnostic performance restricted to patients with biopsy grade group 1-2 disease (n=35).

**Supplemental Figure 1:** Patient selection flowchart

**Supplemental Methods**

Imaging interpretation. Two board-certified nuclear medicine physicians (A.H., S.B.) independently reviewed PSMA PET/CT and two board-certified radiologists (A.C., T.S.) independently reviewed mpMRI, each blinded to clinical data and to one another; prostate-specific antigen was provided. Readers scored extraprostatic extension and seminal vesicle invasion on a 1-5 scale and recorded bilaterality and multifocality, the latter defined as two or more spatially separate foci. Discordances between paired readers were adjudicated by a third reader (I.S. for PET, S.R. for mpMRI) by majority consensus. An independent dual board-certified reader (K.K.), with access to both modalities, performed an integrated read as a secondary analysis (Supplemental Table 1).

Statistical analysis. Ninety-five percent confidence intervals for proportions used the exact (Clopper-Pearson) method, and paired comparisons of sensitivity and specificity between modalities used McNemar tests. Inter-reader agreement for each exclusionary feature was assessed with Cohen's κ between the two PET readers and between the two mpMRI readers, with extraprostatic extension and seminal vesicle invasion dichotomized at a score of ≥3 (Supplemental Table 3). All P values were two-sided, with P < 0.05 considered significant. Analyses used R 4.6.0.

**Supplemental Results**

Sensitivity analysis under a stricter definition. Retaining any bilateral or multifocal disease as exclusionary, in addition to extraprostatic extension, seminal vesicle invasion, and any grade group >3 focus, 90 of 112 patients (80%) were ineligible. Sensitivity was 52% for PSMA PET/CT, 68% for mpMRI, and 80% for the combination (Supplemental Table 4).

Lower-risk subgroup. Among the 35 patients with biopsy grade group 1-2 disease, 22 (63%) were ineligible on whole-mount histopathology. Sensitivity was 59% for PSMA PET/CT, 59% for mpMRI, and 86% for the combination; negative predictive value for combined imaging in this subset was 62%, higher than the 41% observed in the overall cohort (Supplemental Table 5).

Integrated single-reader interpretation. The integrated PET + MRI read by a single dual board-certified reader yielded a sensitivity of 77% and a specificity of 29%, broadly concordant with the rule-based combination, suggesting a practical alternative where multireader workflows are unavailable (Supplemental Table 1).

**Supplemental Table 1:** Diagnostic performance of the integrated PET + MRI read (single dual-board-certified reader)

**Supplemental Table 2:** Distribution of PRIMARY and PI-RADS scores by reader

**Supplemental Table 3:** Inter-reader agreement (Cohen’s κ) between PET readers and between MRI readers

**Supplemental Table 4:** Sensitivity analysis: diagnostic performance under a stricter definition in which any bilateral or multifocal disease (regardless of contralateral grade) was exclusionary, in addition to EPE, SVI, and any focus of GG>3 at prostatectomy

Supplemental Table 5: Diagnostic performance for focal therapy eligibility restricted to patients with biopsy grade group 1-2 disease patients (n=35)

**Supplemental Figure 1:** Patient selection flowchart.

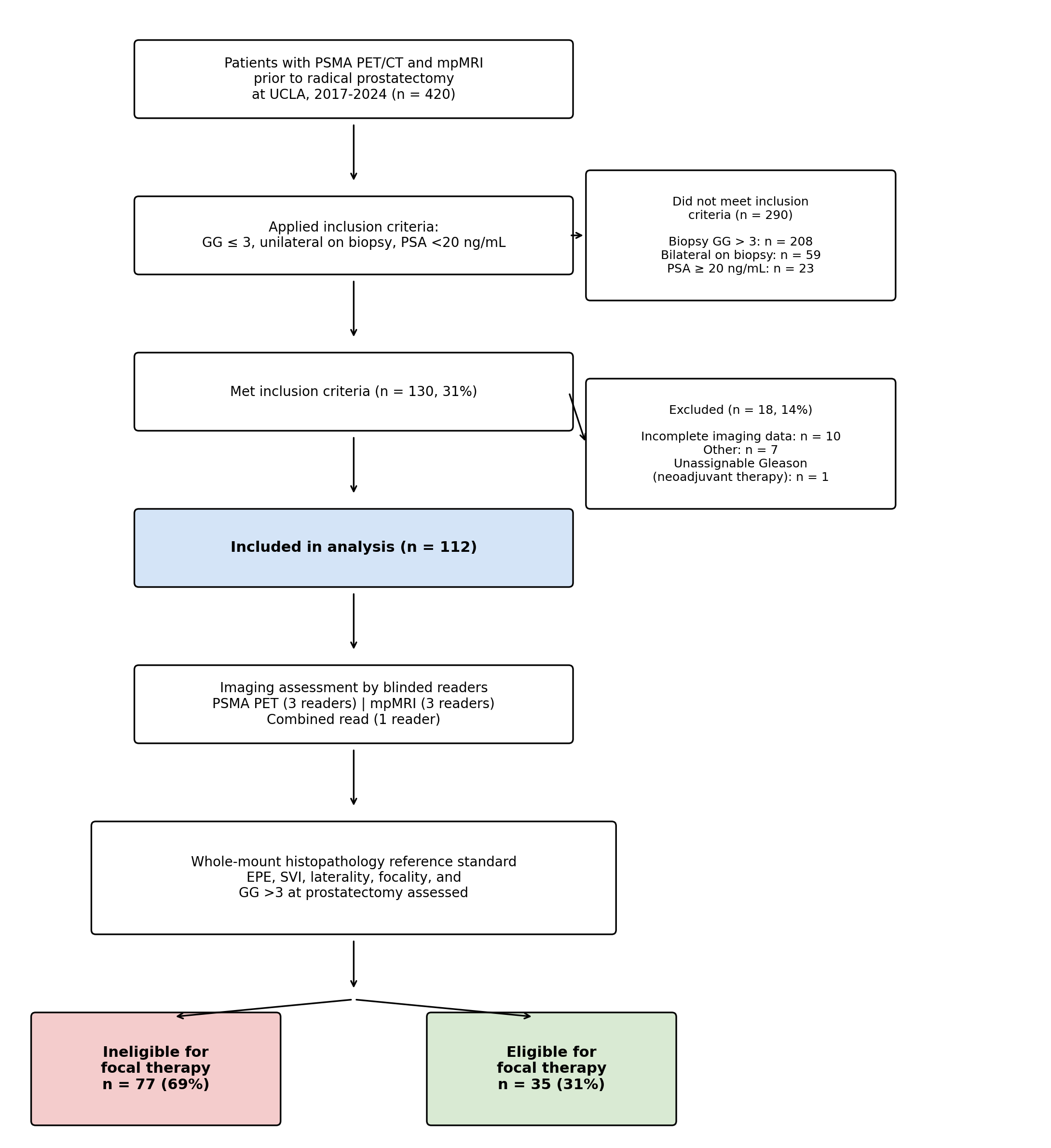

Abbreviations: PSMA, prostate-specific membrane antigen; PET/CT, positron emission tomography/computed tomography; mpMRI, multiparametric magnetic resonance imaging; UCLA, University of California, Los Angeles; GG, Gleason grade group; PSA, prostate-specific antigen; EPE, extraprostatic extension; SVI, seminal vesicle invasion.

**Supplemental Table 1:** Diagnostic performance of the integrated PET + MRI read performed by a single dual-board-certified reader (KK), compared with the rule-based combination and with each single modality.

| **Outcome** | **Modality** | **Sensitivity** | **Specificity** | **PPV** | **NPV** |
| --- | --- | --- | --- | --- | --- |
| **Eligibility (composite)** | PSMA PET/CT | 53 (42-65) | 54 (37-71) | 72 (58-83) | 35 (22-49) |
|  | mpMRI | 70 (59-80) | 49 (31-66) | 75 (63-84) | 42 (27-59) |
|  | PET + MRI (rule-based) | 83 (73-91) | 26 (12-43) | 71 (61-80) | 41 (21-64) |
|  | PET + MRI (integrated) | 77 (66-86) | 29 (15-46) | 70 (59-80) | 36 (19-56) |
| **EPE** | PSMA PET/CT | 32 (20-46) | 90 (79-96) | 74 (52-90) | 60 (49-70) |
|  | mpMRI | 77 (64-88) | 58 (44-70) | 62 (49-74) | 74 (59-86) |
|  | PET + MRI (rule-based) | 83 (70-92) | 54 (41-67) | 61 (48-72) | 78 (62-89) |
|  | PET + MRI (integrated) | 72 (58-83) | 58 (44-70) | 60 (47-72) | 69 (55-82) |
| **SVI** | PSMA PET/CT | 39 (17-64) | 96 (89-99) | 64 (31-89) | 89 (81-94) |
|  | mpMRI | 44 (22-69) | 99 (94-100) | 89 (52-100) | 90 (83-95) |
|  | PET + MRI (rule-based) | 56 (31-78) | 95 (88-98) | 67 (38-88) | 92 (84-96) |
|  | PET + MRI (integrated) | 33 (13-59) | 95 (88-98) | 55 (23-83) | 88 (80-94) |
| **Multifocal disease** | PSMA PET/CT | 35 (22-49) | 73 (60-84) | 53 (35-70) | 56 (45-68) |
|  | mpMRI | 23 (13-37) | 87 (75-94) | 60 (36-81) | 57 (46-67) |
|  | PET + MRI (rule-based) | 44 (30-59) | 67 (53-78) | 49 (34-64) | 55 (43-68) |
|  | PET + MRI (integrated) | 31 (19-45) | 75 (62-85) | 52 (33-70) | 56 (44-67) |
| **Bilateral disease** | PSMA PET/CT | 32 (19-47) | 71 (58-81) | 44 (27-62) | 59 (47-70) |
|  | mpMRI | 23 (12-38) | 86 (75-93) | 55 (32-77) | 61 (50-71) |
|  | PET + MRI (rule-based) | 45 (30-60) | 65 (52-76) | 48 (32-63) | 62 (49-73) |
|  | PET + MRI (integrated) | 38 (25-54) | 65 (52-76) | 44 (28-60) | 59 (47-71) |

Values are shown as percent (95% exact binomial/Clopper-Pearson CI). The integrated read was performed by a single dual-board-certified reader (KK) with simultaneous access to PET and MRI. Rule-based combination = positive if either PSMA PET (majority read) or mpMRI (majority read) was positive. PPV, positive predictive value; NPV, negative predictive value; EPE, extraprostatic extension; SVI, seminal vesicle invasion.

**Supplemental Table 2:** Distribution of PRIMARY and PI-RADS scores by reader (n=112).

| **Score** | **PRIMARY Reader 1 n (%)** | **PRIMARY Reader 2 n (%)** | **PI-RADS Reader 1 n (%)** | **PI-RADS Reader 2 n (%)** | **PI-RADS Reader 3 n (%)** |
| --- | --- | --- | --- | --- | --- |
| 1 / No lesion | 4 (3.5) | 8 (7.1) | 0 (0) | 0 (0) | 0 (0) |
| 2 | 7 (6.2) | 15 (13) | 13 (12) | 6 (5.3) | 3 (2.7) |
| 3 | 15 (13) | 8 (7.1) | 2 (1.8) | 5 (4.4) | 7 (6.2) |
| 4 | 51 (45) | 50 (45) | 53 (47) | 51 (45) | 44 (39) |
| 5 | 35 (31) | 30 (26) | 44 (39) | 50 (45) | 58 (52) |

Per-patient highest PRIMARY and PI-RADS scores recorded by each independent reader. PRIMARY scores were recorded descriptively; PET eligibility determinations were based on expert visual assessment. For Reader 2 of PI-RADS, two scans were rated as “4-equivalent” (grouped with 4) and “5-equivalent” (grouped with 5). One Reader 2 PRIMARY read was incomplete and is not shown.

**Supplemental Table 3:** Inter-reader agreement (Cohen’s κ) between the two PSMA PET readers and between the two mpMRI readers for each exclusionary feature and for the composite focal therapy eligibility endpoint.

| **Feature** | **PET readers (AH vs SB)** | **MRI readers (AC vs TS)** |
| --- | --- | --- |
| Composite eligibility | 0.04 (-0.11-0.20) | 0.38 (0.15-0.62) |
| Extraprostatic extension (EPE) | 0.02 (-0.10-0.14) | 0.52 (0.32-0.71) |
| Seminal vesicle invasion (SVI) | 0.28 (0.03-0.54) | 0.58 (0.35-0.81) |
| Multifocal disease | 0.39 (0.22-0.57) | 0.48 (0.24-0.72) |
| Bilateral disease | 0.48 (0.31-0.65) | 0.60 (0.40-0.80) |

Cohen’s κ for binary feature assessments by reader pair. EPE and SVI were dichotomized at a 1–5 score of ≥3. Composite ineligibility indicates presence of any one exclusionary feature (EPE, SVI, multifocal disease, or bilateral disease) by that reader. 95% CIs computed using Fleiss’ asymptotic SE. Pairs with missing data in either reader were excluded, yielding n=104 (PET) and n=100 (MRI).

**Supplemental Table 4:** Sensitivity analysis: diagnostic performance under a stricter definition in which any bilateral or multifocal disease (regardless of contralateral grade) was exclusionary, in addition to EPE, SVI, and any focus of GG>3 at prostatectomy.

|  | **Sensitivity** | **Specificity** | **PPV** | **NPV** | **p vs PET** | **p vs MRI** |
| --- | --- | --- | --- | --- | --- | --- |
| **PSMA PET/CT** | 52 (41-63) | 55 (32-76) | 82 (70-91) | 22 (12-35) | Ref | 0.029 |
| **mpMRI** | 68 (57-77) | 50 (28-72) | 85 (74-92) | 28 (15-44) | 0.029 | Ref |
| **PET + MRI** | 80 (70-88) | 18 (5-40) | 80 (70-88) | 18 (5-40) | <0.001 | 0.001 |

Under this stricter alternative definition, any bilateral or multifocal disease on whole-mount histopathology was exclusionary regardless of contralateral grade, in addition to EPE, SVI, and any focus of GG>3 at prostatectomy. 90 of 112 patients (80%) were ineligible. P-values in the “p vs PET” and “p vs MRI” columns refer to paired comparisons of sensitivity only, computed from McNemar tests. PPV, positive predictive value; NPV, negative predictive value.

**Supplemental Table 5:** Diagnostic performance for focal therapy eligibility restricted to patients with biopsy grade group 1-2 diseaseble intermediate-risk patients (n=35).

| **Modality** | **Sensitivity** | **Specificity** | **PPV** | **NPV** |
| --- | --- | --- | --- | --- |
| **PSMA PET/CT** | 59 (36-79) | 62 (32-86) | 72 (47-90) | 47 (23-72) |
| **mpMRI** | 59 (36-79) | 77 (46-95) | 81 (54-96) | 53 (29-76) |
| **PET + MRI** | 86 (65-97) | 38 (14-68) | 70 (50-86) | 62 (24-91) |

Eligibility defined per the primary criterion of the main analysis (any extraprostatic extension, seminal vesicle invasion, multifocal/bilateral disease with at least one contralateral or secondary nodule of Grade Group ≥2, or any focus of GG>3 at prostatectomy). The subset comprised 5 biopsy GG1 (low-risk) and 30 biopsy GG2 (favorable intermediate-risk) patients. Of the 35 included, 22 (63%) were ineligible based on whole-mount histopathology. Values for sensitivity, specificity, PPV, and NPV are shown as percent (95% exact binomial/Clopper-Pearson confidence interval). The PET + MRI row reflects a rule-based combination in which a patient was considered imaging-ineligible if either modality was positive by majority read.
